# Evaluation of Statistical Illiteracy in Latin American Clinicians and of the Efficacy of a 10-Hour Course

**DOI:** 10.1101/2021.05.08.21256882

**Authors:** Adrian Soto-Mota, Eduardo Carrillo Maravilla, Jose Luis Cárdenas Fragoso, Óscar Arturo Lozano Cruz, Alfonso Gulías Herrero, Sergio Ponce De Leon Rosales

**Author notes:** **Corresponding author:** Luis Adrian Soto-Mota MD, DPhil., Assistant Professor at the Metabolic Diseases Research Unit of The National Institute of Medical Sciences and Nutrition Salvador Zubirán. Mexico City, Postal code: 14080.

## Abstract

**Introduction:** All medics require statistical interpretation skills to keep up to date with the scientific advances and evidence-based recommendations of their specific field. However, statistical illiteracy among clinicians is a highly prevalent problem with far-reaching consequences. The few available studies that report statistical literacy improvements after educational interventions do not report for how long these benefits last. We measured for the first-time statistical proficiency among Latin-American clinicians with different levels of training and evaluated the efficacy of a 10-hour course at multiple timepoints.

**Methods:** Using an online questionnaire, we evaluated self-perceived statistical proficiency, scientific literature reading habits and statistical literacy (using an adaptation of the Quick Risk Test) across multiple levels of medical training. Separately, we evaluated statistical proficiency among Internal Medicine residents at a tertiary centre in Mexico City immediately before, immediately after and one and two months after a 10-hour statistics course using the same adaptation (allowing for “I don’t know” answers) of the Quick Risk Test. Scores across multiple time points were compared using Friedman’s Test.

**Results:** Data from 392 clinicians from 9 Latin American countries were analyzed. Most clinicians (85%) failed our adaptation of the Quick Risk Test (mean score = 2.6/10, IQR:1.4). The 10-hour course significantly improved the scores of the Internal Medicine Residents (n=16) from 3.8/10, IQR:1.8 to 8.3/10, IQR:1.4 (p<0.01). However, scores dropped after one and two months to 7.7/10, IQR:1.6 and 6.1 / 10, IQR:2.2, respectively.

**Conclusions:** Statistical Illiteracy is highly prevalent among Latin American clinicians. Short-term educational interventions are effective but, their benefits quickly fade away if they are not periodically reinforced. Medical boards and Medical schools need to periodically teach and evaluate statistical proficiency to ameliorate these issues.

## INTRODUCTION

Most medical schools and Medical board recognise the importance of Statistical Skills for practising clinicians^1^. However, evidence shows that even experienced clinicians struggle with assimilating the differences and implications of fundamental statistical concepts such as odds ratio versus absolute risk and sensitivity versus positive post-test probability^2^. Moreover, essential concepts such as absolute risk changes, number needed to treat/screen, intention-to-treat analysis and Bayesian probability are often overlooked when making clinical decisions and when explaining the implications of tests and treatments to patients^3,4^.

The implications of Statistical Illiteracy among clinicians are frequent and range from generating individual ethical problems^5–7^ to health-policy misinformed decisions^8^. Moreover, improving health statistics among medical doctors has been put forward as one of the seven goals for improving health during this century^9^.

Importantly, evidence also suggests that cheap, easy-to-implement and short-term interventions can improve statistical skills among clinicians^10^. In their 2018 study, Jenny, Keller and Gigerenzer^11^ demonstrated that a 90-minute training session in medical statistical literacy improved the performance (from 50% to 90%) in 82% of the participants using a multiple-choice Statistics test. However, it was not evaluated how quickly these improvements fade away after the educational intervention.

In this study, we estimated Statistical Literacy among Latin American clinicians and evaluated the efficacy of a 10-hour Statistics course across multiple timepoints.

## METHODS

This study and its methods were reviewed and approved by y the Ethics Committee of the National Institute of Medical Sciences and Nutrition Salvador Zubirán on September 29th, 2020 (Reg. No. SEN-3516-20-21-1). Data were anonymized, and its collection followed Good Clinical Practice Standards.

### Online survey and Statistics Test

The survey collected information about medical training, medical school and graduation year, self-perceived understanding of the methods section of scientific papers (as a percentage), and the number of scientific papers read per week, and extracurricular statistical training. Email restrictions were placed to ensure respondents were only capable of answering once.

The test was based on the Quick Risk Test^11^ but, to avoid granting points by guessing, was modified to incorporate an “I don’t know” option in all questions. Additionally, it avoided word by word translations and evaluating concepts by directly asking their definitions. Hypothetical cases and examples were used instead. The evaluated concepts were: Sensitivity, Specificity, Positive and Negative Predictive Values, Statistical Power, Sample Size, Statistical Significance, Statistical correlation, Absolute and Relative Risk, Bayesian clinical reasoning and Dependent and Independent probabilities.

Respondents did not receive feedback after answering each question to avoid their early performance influenced their final answers. The survey and questionnaire can be reviewed here at https://forms.gle/fCep4atAhcoG5BKW6

### Characteristics and design of the educational intervention

A 10-hour course was dived into ten one-hour weekly sessions to review each one of the concepts evaluated by the test. All sessions were recorded and available for review during the 10 weeks the course lasted.

This course was summarized into a 3-hour long 10 session videos now freely available at: https://www.youtube.com/watch?v=cdEX8AdEU6Y&list=PLoieIsf7siGMTkICPbkvgD1hyZpbVwd0H

### Evaluation of the efficacy of the course

Internal Medicine residents answered the aforementioned survey and test before the course, immediately after the last session, one month after the course and two months after the course. Lecture recordings were unavailable after the course ended to avoid biasing the follow-up evaluations.

The same questions were used for all evaluations except for the very last one in which, different cases evaluated the same Statistical concepts.

### Statistical Analysis

Since scores were not normally distributed, we compared them using Friedman’s Test using the R function “friedman.test” from the R package “stats” version 3.6.2. Normality was evaluated using Shapiro-Wilk tests using the function “shapiro.test” in the same R package.

## RESULTS

### Survey responses and Statistical Literacy results among Latin American clinicians

A total of 403 responses were collected, however, 11 were discarded due to having incomplete data. In total 392 from 9 different countries and 53 different medical schools were included in the analysis. Most respondents (82%) were Mexican. Scores were not significantly different across different levels of medical training. Table 1 summarizes their answers in the survey and the overall performance in the test. Table 2 summarizes the percentage of correct answers in every statistical concept we evaluated.

**TABLE 1.** Participants educational background and survey results.

| **n= 392** | **(IQR / standard deviation)** |
| --- | --- |
| Medical training level |  |
| Undergraduate | 26.3 % |
| General Practitioner | 26.0 % |
| Resident | 11.3 % |
| Graduated Specialist | 36.4 % |
| Years from medical graduation | 6 (+/- 3) |
| Scientific papers read per week* | 3 (+/- 2) |
| Self-perceived level of Methodological understanding. | 55 (+/- 17) |
| Extra-curricular statistical training | 21% |
| Test score (on a scale of 10) | 2.6 (+/-1.4) |
| Test scores by medical training level |  |
| Undergraduate | 2.7/10 (+/- 1.5) |
| General Practitioner | 2.2/10 (+/- 1.8) |
| Resident | 3.4/10 (+/- 1.3) |
| Graduated Specialist | 2.9/10 (+/- 1.4) |
| Overall tests score | 2.6/10 (+/- 1.4) |

**TABLE 2.** Performance in every evaluated concept.

| Statistical concept | % of correct answers | % “I don’t know” |
| --- | --- | --- |
| Sensitivity and Specificity | 24 % | 11 % |
| Positive and Negative Predictive Values | 63 % & 15 % | 3% & 12% |
| Statistical Power | 32 % | 16% |
| Sample Size | 19 % | 10% |
| Statistical Significance | 37 % | 31% |
| Absolute and Relative Risk | 18% | 21% |
| Bayesian clinical reasoning | 45% | 5% |
| Statistical correlation | 31% | 26% |
| Dependent and Independent probabilities | 14 % | 5% |

### Evaluation of the efficacy of the course

Internal Medicine residents at the National Institute of Medical Sciences and Nutrition Salvador Zubirán voluntarily attended or listened to the recorded lectures at their own pace and answered the Statistics tests at the already mentioned time points. Tests were self-paced, unsupervised and were open for 5 days during each time point. Email restrictions were placed to allow no more than one answer per resident.

Only those who answered all tests (n=16 out of 42) were included in the analysis. Figure 1 shows the scores results and their distributions across the evaluated time points. Timepoints were statistically different from baseline when compared with Friedman’s Test (p<0.01).

**FIGURE 1.**
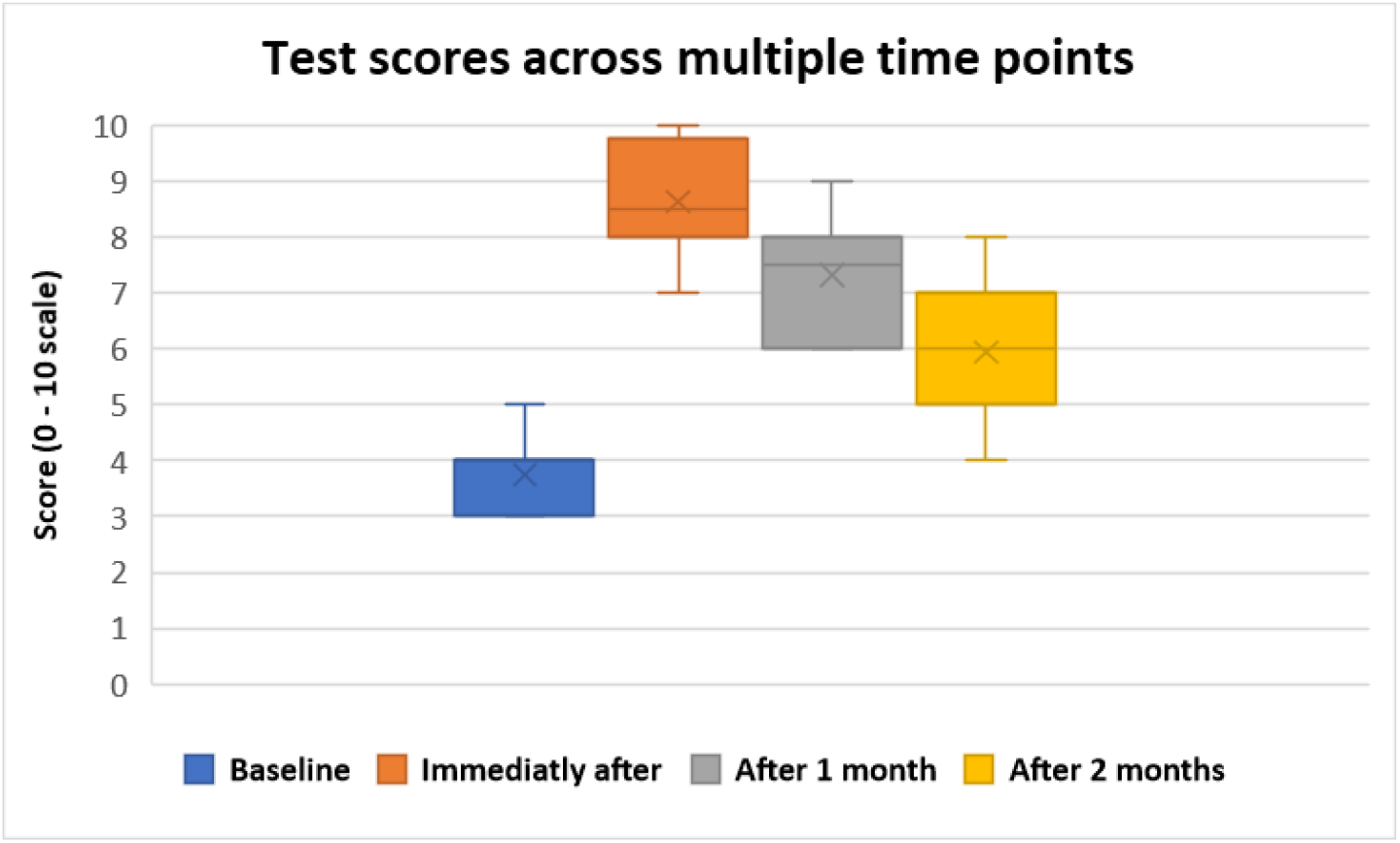
Test score across multiple time points. Timepoints were statistically different from baseline when compared with Friedman’s Test (p<0.01). n=16.

Further, follow up was stopped because the resident’s academic year ended in March 2021.

## DISCUSSION

To our knowledge, this is the first attempt at estimating Statistical illiteracy among Latin American Clinicians. Despite having found comparable statistical proficiency scores to those in other countries^11^, this population merits being analysed separatedly because most educational tools that address this problem are only available in English^12,13^ and English proficiency is not mandatory for practicing Medicine in all Latin American countries.

In contrast with Jenny, Keller and Gigerenzer study^11^, we allowed for respondents to admit they did not know an answer to the questions and measured for up to two months the lasting effects of the educational intervention we tried. The fact that scores quickly and significantly drop after a few weeks of having finished the course highlight the importance of continuous teaching and periodical evaluation. The finding that it is infrequent that clinicians recognise they do not know and the discrepancy between their self-perceived statistical skills and their tests scores suggest clinicians overestimate their statistical proficiency. Teaching clinicians to identify when they lack enough information or how to avoid cognitive biases should be emphasized when designing educational tools.

Some questions evaluated the same concepts by evaluating theoretical knowledge (i.e., Which of the following factors influences the the positive predictive value of a test?) while others by presenting practical scenarios (i.e., How does the positive predictive value of an Influenza rapid test changes over the year?). Interestingly, clinicians performed better at theoretical questions (63% right answers) than with practical ones (15% right answers). Thus, it is likely than emphasizing practical interpretation over theoretical knowledge would yield better results when designing educational resources for improving statistical literacy.

The main limitations of our study are inherent to the nature of self-reported, online-based studies. Also, Mexican clinicians were overrepresented in our sample. Nonetheless, the large number of participants and consistency of the results make it unlikely that more controlled methods would yield very different results.

An additional limitation for extrapolating the utility of these short-term interventions comes from the fact there is not a consensus about which specific statistical skills are necessary for all physicians. Moreover, different types of specialists would likely require developing and preserving different skills. For example, clinical trials are more frequent in Internal Medicine Journals than in Forensic Medicine ones.

Nonetheless, since it is not possible to practise Evidence-Based Medicine if clinicians cannot understand scientific evidence, further research is much needed to help guide future educational strategies and policies that help reduce the educational, ethical and economical impact Statistical Illiteracy has on everyday medical practice.

## CONCLUSION

Similarly, to other populations, most Latin American clinicians struggle with statistical concepts that are essential for correctly interpreting emerging evidence. Short-term educational interventions can improve statistical skills; however, these improvements quickly fade away if they are not reinforced. Our results highlight the need to periodically teach and evaluate statistical proficiency by Medical schools and Medical boards.

## Data Availability

Data for research purposes will be shared upon request to the corresponding author. Dr Adrian Soto-Mota is the guarantor of the integrity of this work.

## ACKNOWLEDGMENTS

The authors want to thank Dr Ricardo Macias for his help in distributing the online survey.

## FUNDING, DATA SHARING AND CONFLICT OF INTEREST DISCLOSURES

This study did not receive funding. The authors declare they do not have conflicts of interest to disclose. Data for research purposes will be shared upon request to the corresponding author. Dr Adrian Soto-Mota is the guarantor of the integrity of this work.

## Notes

### Competing Interest Statement

The authors have declared no competing interest.

### Funding Statement

This study received no funding.

### Author Declarations

This study and its methods were reviewed and approved by y the Ethics Committee of the National Institute of Medical Sciences and Nutrition Salvador Zubiran on September 29th, 2020 (Reg. No. SEN-3516-20-21-1). Data were anonymized, and its collection followed Good Clinical Practice Standards.

## REFERENCES

1. Johnson T v., Abbasi A, Schoenberg ED, et al. Numeracy among trainees: Are we preparing physicians for evidence-based medicine? Journal of Surgical Education. 2014;71(2):211–215. Doi:10.1016/j.jsurg.2013.07.013

2. Anderson BL, Williams S, Schulkin J. Statistical Literacy of Obstetrics-Gynecology Residents. Journal of Graduate Medical Education. 2013;5(2):272–275. Doi:10.4300/jgme-d-12-00161.1

3. Naylor CD, Chen E, Strauss B. Measured enthusiasm: Does the method of reporting trial results alter perceptions of therapeutic effectiveness? Annals of Internal Medicine. 1992;117(11):916–921. Doi:10.7326/0003-4819-117-11-916

4. Whiting PF, Davenport C, Jameson C, et al. How well do health professionals interpret diagnostic information? A systematic review. BMJ Open. 2015;5(7). Doi:10.1136/bmjopen-2015-008155

5. Entwistle VA, Carter SM, Cribb A, McCaffery K. Supporting patient autonomy: The importance of clinician-patient relationships. Journal of General Internal Medicine. 2010;25(7):741–745. Doi:10.1007/s11606-010-1292-2

6. Sedrakyan A, Shih C. Improving depiction of benefits and harms: Analyses of studies of well-known therapeutics and review of high-impact medical journals. Medical Care. 2007;45(10 SUPPL. 2): S23–8. Doi:10.1097/MLR.0b013e3180642f69

7. Wegwarth O, Gigerenzer G. The barrier to informed choice in cancer screening: Statistical illiteracy in physicians and patients. In: Recent Results in Cancer Research. Vol 210. Springer New York LLC; 2018.207–221. Doi:10.1007/978-3-319-64310-6_13

8. Iacobucci G. Conservative conference: May announces new cancer strategy to boost survival rates. BMJ (Clinical research ed). 2018;363:k4198. Doi:10.1136/bmj.k4198

9. Gigerenzer G, Gray JAM. Launching the Century of the Patient. (Gigerenzer G, Gray JAM, eds.). The MIT Press; 2011. Doi:10.7551/mitpress/9780262016032.001.0001

10. Garcia-Retamero R, Cokely ET, Wicki B, Joeris A. Improving risk literacy in surgeons. Patient Education and Counseling. 2016;99(7):1156–1161. Doi: 10.1016/j.pec.2016.01.013

11. Jenny MA, Keller N, Gigerenzer G. Assessing minimal medical statistical literacy using the Quick Risk Test: A prospective observational study in Germany. BMJ Open. 2018;8(8):e020847. Doi:10.1136/bmjopen-2017-020847

12. Understanding Medical Research: Your Facebook Friend is Wrong | Coursera. Accessed May 4, 2021. https://www.coursera.org/learn/medical research.

13. Stanford Medical Statistics Certificate | Stanford Online. Accessed May 4, 2021. https://online.stanford.edu/programs/stanford-medical-statistics-certificate

